# Cerebrovascular Lesion Loads and Accelerated Brain Ageing: Insights into the Cognitive Spectrum

**DOI:** 10.1101/2023.10.04.23296558

**Authors:** Iman Beheshti, Olivier Potvin, Mahsa Dadar, Simon Duchesne, the CCNA Group

## Abstract

**INTRODUCTION:** White matter hyperintensities (WMHs) and cerebral microbleeds are widespread among aging population and linked with cognitive deficits in mild cognitive impairment (MCI), vascular MCI (V-MCI), and Alzheimer’s disease without (AD) or with a vascular component (V-AD). In this study, we aimed to investigate the association between brain age, which reflects global brain health, and cerebrovascular lesion load in the context of pathological aging in diverse forms of clinically-defined neurodegenerative conditions.

**METHODS:** We computed brain-predicted age difference (brain-PAD: predicted brain age minus chronological age) in the Comprehensive Assessment of Neurodegeneration and Dementia cohort of the Canadian Consortium on Neurodegeneration in Aging including 70 cognitively intact elderly (CIE), 173 MCI, 88 V-MCI, 50 AD, and 47 V-AD using T1-weighted magnetic resonance imaging (MRI) scans. We used a well-established automated methodology that leveraged fluid attenuated inversion recovery MRIs for precise quantification of WMH burden. Additionally, cerebral microbleeds were detected utilizing a validated segmentation tool based on the ResNet50 network, utilizing routine T1-weighted, T2-weighted, and T2* MRI scans.

**RESULTS:** The mean brain-PAD in the CIE cohort was around zero, whereas the four categories showed a significantly higher mean brain-PAD compared to CIE. The brain-PAD was significantly correlated with WMHs in all groups.

**DISCUSSION:** WMHs were associated with faster brain ageing and should be considered as a risk factor which imperils brain health in aging and exacerbate brain abnormalities in the context of neurodegeneration of presumed AD origin. Our findings underscore the significance of novel research endeavors aimed at elucidating the etiology, prevention, and treatment of WMH in the area of brain ageing.

**Highlights:**

- We assessed the association between brain-PAD and cerebrovascular lesion loads in aging and AD.
- There were noticeably links between brain-PAD and WMH loads.
- The structure of the ageing brain is associated with WMHs.
- WMH needs to be taken into account as a risk factor that increase the brain age in aging and AD.

## 1. INTRODUCTION

Assessing brain health status using machine learning models is a topic of increasing interest, with a number of useful applications being proposed such as predicting future decline trajectories in neurodegeneration [1]. A recent addition consists in predicting a “brain age” metric as an indicator of cerebral health[2–4], whereby observable characteristics from neuroimaging – e.g., cortical thickness as measured on magnetic resonance imaging (MRI) – are used as dependent variables in an estimation framework of a group of individual’s chronological age [5]. This allows the generation of a brain age estimate for any new individual based on similar characteristics, with any discrepancy between chronological and predicted brain ages being seen as a departure from the norm defined by the initial training set. To date, a wide range of studies have used this brain age estimation paradigm to assess the impact of different neurological diseases on brain status [2, 6–8], or elucidate the effect of metabolic factors (e.g., diabetes) and mental health on healthy brain aging [9].

In the area of Alzheimer’s disease (AD), the brain age paradigm has been used to uncover its association with traditional neuropsychological screening tools [10], predict the conversion of mild cognitive impairment (MCI) to probable AD [7], and study the trajectory of metabolism along the cognitive impairment spectrum [6]. In fact, cortical thickness-based brain age has been shown to be a stronger predictor of cognitive impairment than chronological age [11]. However, these previous studies have only focused on the spectrum of dementia from probable AD. Clinical-pathological studies have shown however that, while being the most frequent pathology, the incidence of “pure” AD is low (around 6% of individuals with a major neurocognitive disorder) [12]. In fact, more than three-quarters of individuals exhibit two or more pathologies at death, the most prevalent combination being AD and vascular pathology [13]. Rather than the exception, mixed presentations are therefore the norm and should be studied together whenever possible. The clinical diagnostic of vascular MCI (V-MCI) and mixed vascular-AD dementia (V-AD) relies on a series of clinical criteria (see section 2.2.2) as well as on the presence of cerebrovascular lesions on computer tomography or MRI scans, such as microbleeds and white matter hyperintensities (WMH). The latter appear on T2-weighted and fluid attenuated inversion recovery (FLAIR) scans and are typically seen in the aging population [14]. The prevalence of WMH is 10-20% for people in their 60s, reaching 100% in people over the age of 90 [15]. In the aging population, WMHs can contribute to a higher rate of brain atrophy in beyond-normal brain ageing, particularly in regions related to AD [16]. Recent studies have revealed a pervasive presence of WMH among patients with diverse neurodegenerative disorders, encompassing conditions such as MCI and AD; those with an associated vascular component, V-MCI and V-AD; cognitively intact elderly with Parkinson’s disease; cognitively impaired Parkinson’s disease; frontotemporal dementia; Lewy body dementia; and mixed dementias [14]. Remarkably, the manifestation of WMHs has been associated with a broad spectrum of histological alterations, including demyelination and axonal loss, diminished glial density and atrophy, cortical thinning and cerebral atrophy, endothelial and immune activation, ischemic damage, as well as hypoxia and hypoperfusion[17, 18]. In the context of AD, it has been shown that a heavy burden of WMH can lead to an elevated risk of dementia due to AD and a faster progression from intact cognition to MCI [19]. Likewise, a similar narrative unfolds regarding the burden of microbleeds, wherein their presence can have a detrimental effect on cognitive functioning and can make individuals more vulnerable to dementia [20, 21].

In this study, we aimed to explore brain aging in various populations along both spectra of cognition and cerebrovascular disease, as evidenced by the presence of a cerebrovascular lesion load that includes both WMHs and microbleeds. In view of the relevant literature, we hypothesized that cerebrovascular lesion load would be associated with an elevated brain age not only in the context of AD, but also among cognitively healthy older adults. To this end, we estimated cortical morphometric-based brain age in a large group of participants from the Comprehensive Assessment of Neurodegeneration and Dementia (COMPASS-ND) study that included cognitively intact elderly (CIE), individuals with MCI and probable AD, but also participants with V-MCI and V-AD, with specific attention to the associations between brain age and cerebrovascular lesions.

## 2. MATERIAL AND METHODS

### 2.1. Ethical agreement

Ethical agreements were obtained at all respective sites. Written informed consent was obtained from all participants.

### 2.2. Participants

#### 2.2.1 Training set

The data used to train the brain age estimate model were obtained from CIE participants enrolled in the Open Access Series of Imaging Studies (OASIS), Alzheimer’s Disease Neuroimaging Initiative (ADNI), Banner Alzheimer’s Institute (BAI), and Alzheimer’s Disease Repository Without Borders (ARWIBO) studies.

ADNI (adni.loni.usc.edu) was launched in 2003 as a public-private partnership led by Principal Investigator Michael W. Weiner, MD. The primary goal of ADNI has been to test whether serial MRI, positron emission tomography, other biological markers, and clinical and neuropsychological assessment can be combined to measure the progression of MCI and early AD. ADNI was carried out with the goal of recruiting 800 adults aged from 55 to 90 years and consists of approximately 200 cognitively normal patients, 400 patients with MCI, and 200 patients with AD (http://adni.loni.usc.edu/wp-content/uploads/2010/09/ADNI_GeneralProceduresManual.pdf).

#### 2.2.2 Test set

The data on our test set participants were collected in the Canada-wide multi-center, prospective, longitudinal COMPASS-ND cohort study of the Canadian Consortium for Neurodegeneration and Aging (CCNA; http://ccna.dev.simalam.ca/compass-nd-study/) [22], a national initiative to catalyze research on dementia. The overall study design and methods have been published previously [22]. The study is registered on clinicaltrials.gov (NCT03402919). COMPASS-ND includes deeply phenotyped participants with various forms of dementia and mild memory loss or concerns, along with CIE. Clinical diagnoses were determined by participating clinicians based on longitudinal clinical, screening, and MRI findings (i.e., diagnosis reappraisal was performed using information from recruitment assessment, screening visit, clinical visit with physician input, and MRI). In particular, criteria for V-MCI were derived from consensus criteria from the American Heart Association [23] and International Society for Vascular Cognitive and Behavioral Disorders [24]. V-MCI participants were required to be age ≥60, have MCI according to National Institute on Aging-Alzheimer’s Association criteria [25], not have a prior history of clinical stroke, and to have evidence of cerebrovascular disease on brain MRI defined as two or more supratentorial infarcts (i.e., excluding brainstem or cerebellar infarcts) or beginning confluent or confluent WMH. Criteria for mixed dementia were adapted from National Institute on Aging-Alzheimer’s Association criteria for dementia due to AD [26] and required that a non-AD cause of dementia should additionally be present.

This cohort included COMPASS-ND participants for whom T1-weighted, T2-weighted, T2*, and FLAIR MRI were obtained. Of note, these data were completely independent from the data used for training the brain age algorithm.

### 2.3 Image acquisition and processing

The acquisition of COMPASS-ND MRIs was done according to the Canadian Dementia Imaging Protocol (CDIP; https://www.cdip-pcid.ca) [27]. T1-weighted images were used to extract cortical thickness measurements from which brain age was derived. To this end, we utilized the *FreeSurfer* version 6.0 segmentation software (http://surfer.nmr.mgh.harvard.edu) and the Desikan-Killiany-Touville atlas [28] to extract neocortical measurements (i.e. surface, volume and thickness extracted from *aparc.DKTatlas* files). Each brain segmentation was visually inspected through at least 20 evenly distributed coronal sections. This procedure was applied to both training and test sets.

WMH load was extracted from T2w-FLAIR images from the COMPASS-ND study. We used a previously validated technique which segments WMHs in native FLAIR space and generates total WMH loads [29], publicly available at: http://nist.mni.mcgill.ca/white-matter-hyperintensities/. For each participant, WMH load was quantified as the volume of voxels that have been categorized as WMH in the standard space, adjusted for head size. The quality of WMH processing and segmentation was visually assessed for quality by one expert (M.D.), resulting in the exclusion of nine cases from the total of 976. A logarithmic transformation was implemented on WMH volumes to normalize their distribution.

The identification of cerebral microbleeds was accomplished using a validated segmentation tool that works based on the ResNet50 network and routine T1-weighted, T2-weighted, and T2* MRI scans, as described in [30]. Visual assessment of the quality of T1, T2, and T2* MRI scans was conducted by the CCNA neuroimaging team prior to applying our cerebral microbleed segmentation tool. In every participant, the total number of cerebral microbleeds was extracted and then log-transformed to normalize the overall distribution. Due to their missing T2*-weighted MRI, twenty-two participants were excluded from cerebral microbleed analysis: five with CIE, seven with MCI, five with AD, four with V-MCI, and one with V-AD.

### 2.4 Brain age estimation model

A standard linear support vector regression algorithm conducted in MATLAB (i.e., “fitrsvm” function, kernel: linear) was used to estimate brain age. Chronological age was considered the dependent variable, and anatomical measurements extracted from *FreeSurfer* segmentation along with sex and total intracranial volume were the independent variables, in total n=188 features per individual. Our brain age prediction model was based on cortical mean thickness, volume, and surface measurements, omitting any white matter related features. First, we assessed the accuracy of the prediction model on the training data set (N = 1627, mean age = 67.75±9.53 years, 56% females) through a 10-fold cross-validation strategy. The prediction accuracy was measured on the basis of the coefficient of determination (R^2^) between chronological and estimated age, the mean absolute error (MAE), and root mean square error (RMSE). The brain-PAD (i.e., predicted brain age minus real age) was also calculated. Bias adjustment was applied to remove the age-dependency on the predicted values [31] (https://github.com/medicslab/Bias_Correction). Next, the final prediction model was developed with the entire training set (*N* =1627). Of note, a positive brain-PAD (strictly speaking, cortical brain-PAD) stands for older-appearing cortices (i.e., estimated age > chronological age), whereas a negative brain-PAD value stands for younger-appearing cortices (i.e., estimated age < chronological age).

### 2.5 Statistical analysis

The brain age prediction model was applied to COMPASS-ND participants. The mean brain-PAD and WMHs for each group (CIE, MCI, AD, V-MCI, V-AD) were examined using an analysis of covariance (ANCOVA) whilst adjusting for sex and age. The p-values were adjusted using Bonferroni correction. Partial correlation tests were utilized to assess the associations between brain-PAD and WMH, as well as between brain-PAD and microbleed loads, while controlling for age and sex as covariates. For all statistical tests, *P* < 0.05 was considered as significant.

## 3. RESULTS

### 3.1 Demographics

The training set was composed of n = 1627 CIE (mean age ± sd: 67.7 ± 9.5, age range: 50–94, 915 females). The test set was composed of 70 CIE, 173 MCI, 50 V-MCI, 88 AD, and 47 V-AD participants. Table 1 shows demographics for our different groups. Of note, mean age was different across diagnostic groups.

**Table 1.** Clinical demographics, WMH and brain-PAD by diagnosis.

| Cohort | N | Female (%) | Real age (yrs) | WMH load | Microbleed count | Brain-PAD (yrs), |
| --- | --- | --- | --- | --- | --- | --- |
| CIE | 70 | 78% | 69.8 ± 6.6 | 1.87±0.65<sup>a</sup> | 2.83±0.73<sup>b</sup> | -0.54 ± 4.75 |
| MCI | 173 | 45% | 71.8 ± 6.6 | 1.98±0.56<sup>c</sup> | 2.98±0.64<sup>d</sup> | 0.98 ± 5.75 |
| AD | 50 | 36% | 73.9 ± 8.2 | 2.24±0.47 | 2.86±0.77<sup>e</sup> | 7.23 ± 7.34 |
| V-MCI | 88 | 34% | 76.1 ± 5.5 | 3.11±0.57<sup>f</sup> | 3.11±0.6<sup>f</sup> | 3.25 ± 5.89 |
| V-AD | 47 | 49% | 77.7 ± 6.3 | 3.20±0.56<sup>f</sup> | 3.20±0.70<sup>e</sup> | 7.61 ± 5.95 |

CIE: cognitively intact elderly, MCI: mild cognitive impairment, AD: Alzheimer's dementia, V-MCI: vascular MCI, V-AD: vascular AD. All variables are presented based on the mean ± standard deviation. Note: WMH loads and microbleed counts were log-transformed.
<sup>a</sup>Data missing in two participants.
<sup>b</sup>Data missing in five participants.
<sup>c</sup>Data missing in thirteen participants.
<sup>d</sup>Data missing in seven participants.
<sup>e</sup>Data missing in four participants.
<sup>f</sup>Data missing in three participants.

### 3.2 Brain age estimation performance on training set

Our prediction model showed an outstanding performance on the training set (N = 1627) followed by 10-fold cross-validation (R^2^ = 0.83, MAE =3.3 years, RMSE = 4.4 years, mean brain-PAD = 0 ± 4.3 years).

### 3.3 Brain age estimation on the test set

As per the initial aim of this study, we computed brain-PAD among five categories of participants. The mean brain-PAD values are shown in Table 1 and Figure 1. There was a significant difference in brain-PAD [F (4,421) = 17, *P* < 0.001, ANCOVA test] among groups, whilst adjusting for sex and chronological age. All categories of patients exhibited a significantly higher mean brain-PAD than the CIE group (P < 0.001), except for the MCI cohort (*P* = 0.38). The V-AD cohort had the highest brain-PAD. An ANCOVA revealed no main effect of sex (F (1, 421) = 0.83, *P* = .36) and the predicted main effect of chronological age (F (1, 421) = 0.71, *P* = 0.39). Post hoc pairwise group comparison based on the ANCOVA test (Fig 3) showed statistically significant differences (*P* < 0.001) in terms of brain-PAD between pair groups, except for CIE vs. MCI, V-AD vs. V-MCI and V-AD vs. AD (*P* > 0.05).

**Figure 1:**
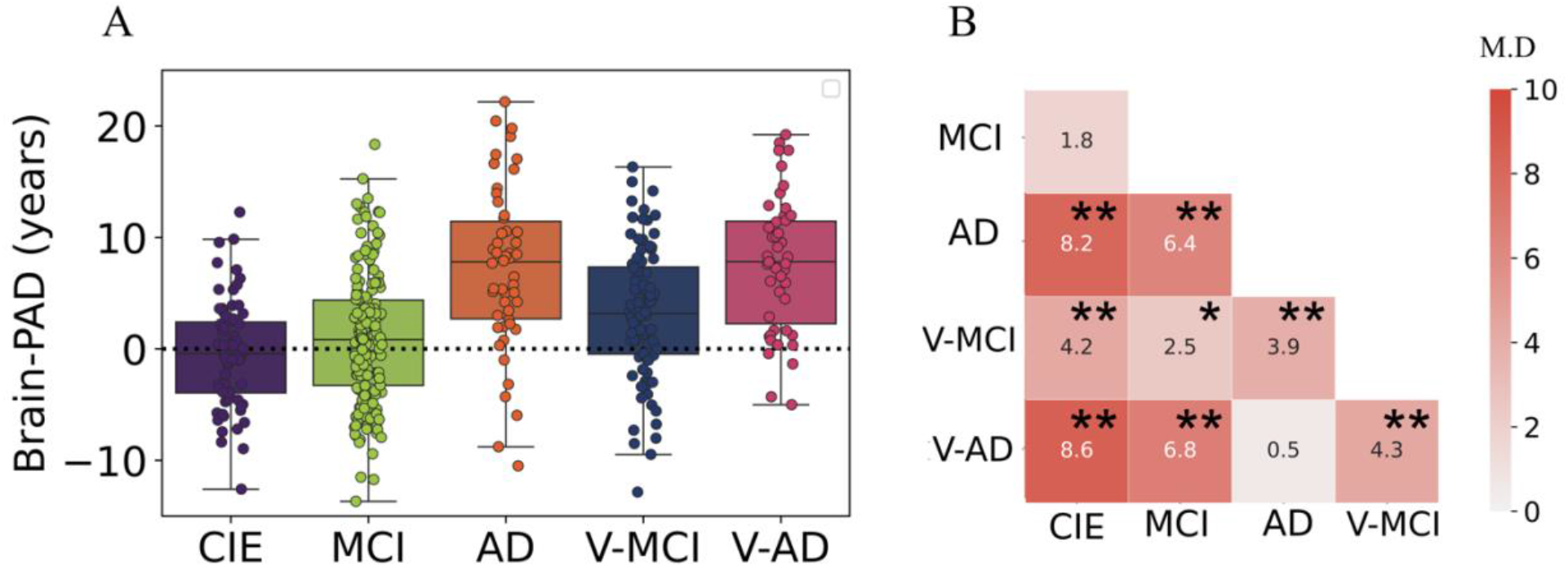
A) Boxplots depicting the brain age delta values among different cohorts. B) Pairwise comparisons. Color bar and numbers inside the boxes stand for absolute of mean difference values between pair groups. CIE: cognitively intact elderly, MCI: mild cognitive impairment, AD: Alzheimer’s dementia, V-MCI: vascular MCI (V-MCI), (AD), V-AD: vascular AD. * *P* < 0.05, ** *P* < 0.001. *p*-value is adjusted using Bonferroni correction.

### 3.4 WMH loads

Table 1 summarizes WMH loads (log-transformed) by diagnostic category, whereas Fig. 2 shows respective boxplots as well as pairwise comparisons. As could be expected, there was a significant difference in WMH loads [F (4,400) = 89, *P* < 0.001, ANCOVA test] between groups, whilst adjusting for sex and age. The ANCOVA revealed an insignificant main effect of sex (F (1, 400) = 1.43, P = 0.23) on WMH loads among the five groups, but age was significant (F (1, 400) = 88, P < 0.001). Unsurprisingly, both V-MCI and V-AD showed a significantly higher log-transformed WMH load compared to non-vascular groups (i.e., CIE, MCI and AD) in terms of WMH loads by ANCOVA pairwise comparison (*P* < 0.001). However, there were no pairwise differences between CIE vs. MCI and V-MCI vs. V-AD (*P* > 0.05).

**Figure 2:**
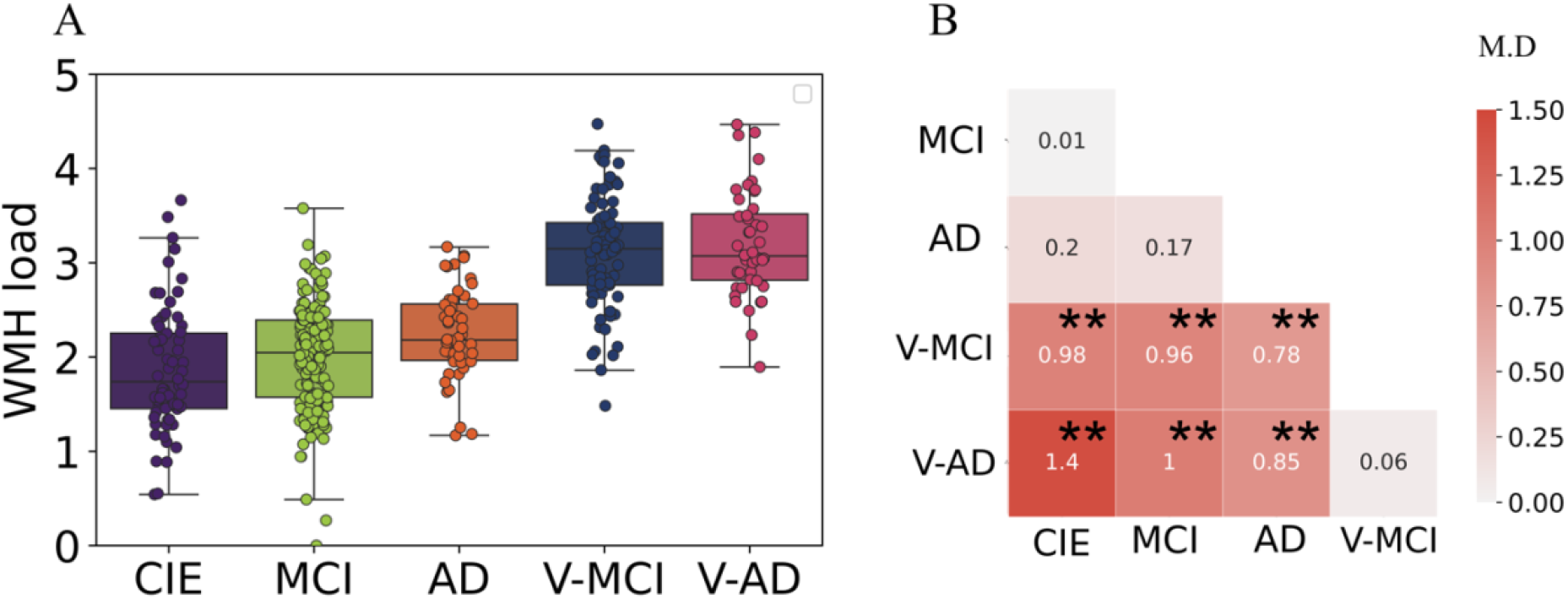
A) Boxplots depicting the WMH loads among different cohorts. B) Pairwise comparisons. Color bar and numbers inside the boxes stand for absolute of mean difference (M.D.) values between pair groups. CIE: cognitively intact elderly, MCI: mild cognitive impairment, AD: Alzheimer’s dementia, V-MCI: vascular MCI (V-MCI), (AD), V-AD: vascular AD. * *P* < 0.05, ** P < 0.001. *p*-value is adjusted using Bonferroni correction. WMH loads were log transformed.

### 3.5 Microbleed counts

The microbleed counts, which have been log-transformed, are presented in Table 1 according to diagnostic category. Figure 3 illustrates the corresponding boxplots and pairwise comparisons. There was a significant difference [F (4,398) = 6, *P* < 0.001, ANCOVA test] between groups in terms of microbleed counts, whilst adjusting for sex and age. Based on the results of an ANCOVA, it was found that there was a significant main effect of sex (F (1, 398) = 17, P < 0.001) and age (F (1, 398) = 4, P = 0.038) on the microbleed counts among the five groups. Based on the results of ANCOVA pairwise comparison, a notable differentiation was observed between AD and V-MCI with regards to microbleed counts (P = 0.016). Conversely, the remaining pair comparisons did not yield significant outcomes (P > 0.05).

**Figure 3:**
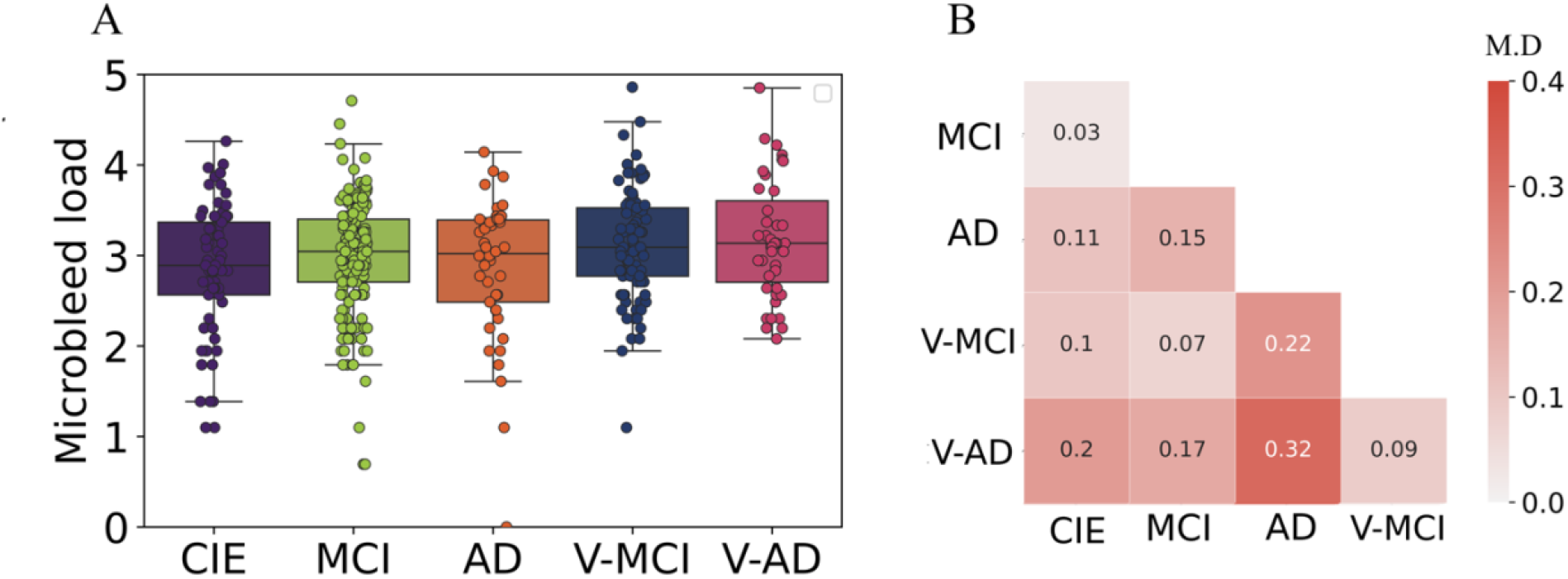
Boxplots showing the microbleed counts in different cohorts. B) Pairwise comparisons. Color bar and numbers inside the boxes stand for absolute of mean difference (M.D.) values between pair groups. CIE: cognitively intact elderly, MCI: mild cognitive impairment, AD: Alzheimer’s dementia, V-MCI: vascular MCI (V-MCI), (AD), V-AD: vascular AD. * *P* < 0.05, ** P < 0.001. *p*-value is adjusted using Bonferroni correction. Microbleed counts were log transformed.

### 3.6 Association between brain-PAD and WMH

Figure 4 shows the association between brain-PAD and WMH loads in the five categories of participants. Brain-PAD and WMH loads demonstrated a significant and positive correlation in the MCI cohort as well as all cohorts combined (r = 0.36, *P* < 0.001), while in other cohorts this association was found to be marginally insignificant (*P* > 0.05).

**Figure 4:**
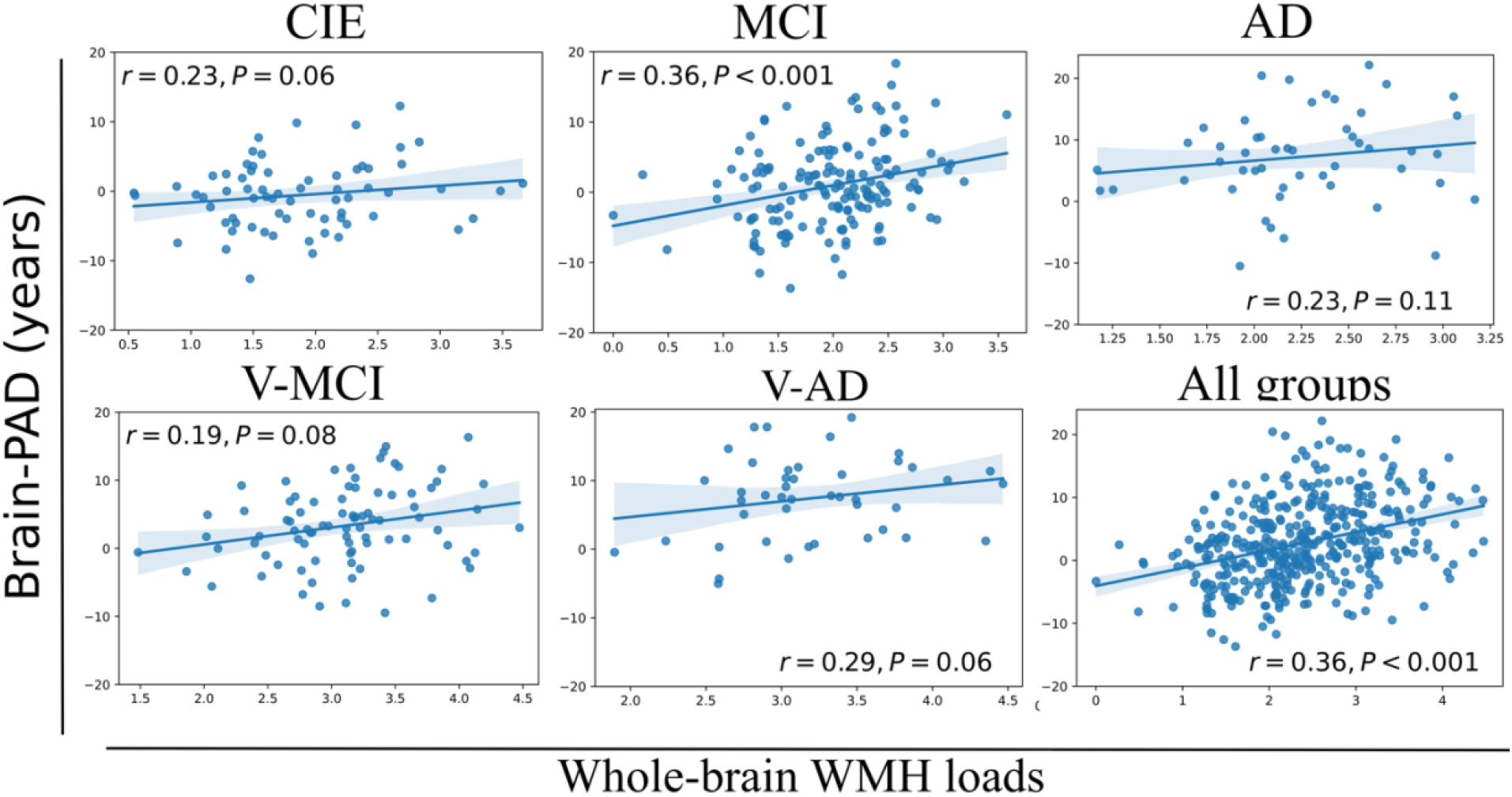
The association between brain-PAD values and whole-brain WMH loads in each cohort, as well as in all cohorts. The correlation analysis was conducted using a partial correlation test while controlling for age and sex. WMH loads were log-transformed.

### 3.7 Association between brain-PAD and microbleed counts

Figure 5 illustrates the correlation between brain-PAD and the number of microbleeds across the five participant groups. There was no statistically significant correlation observed between brain-PAD and microbleed counts in all cohorts.

**Figure 5:**
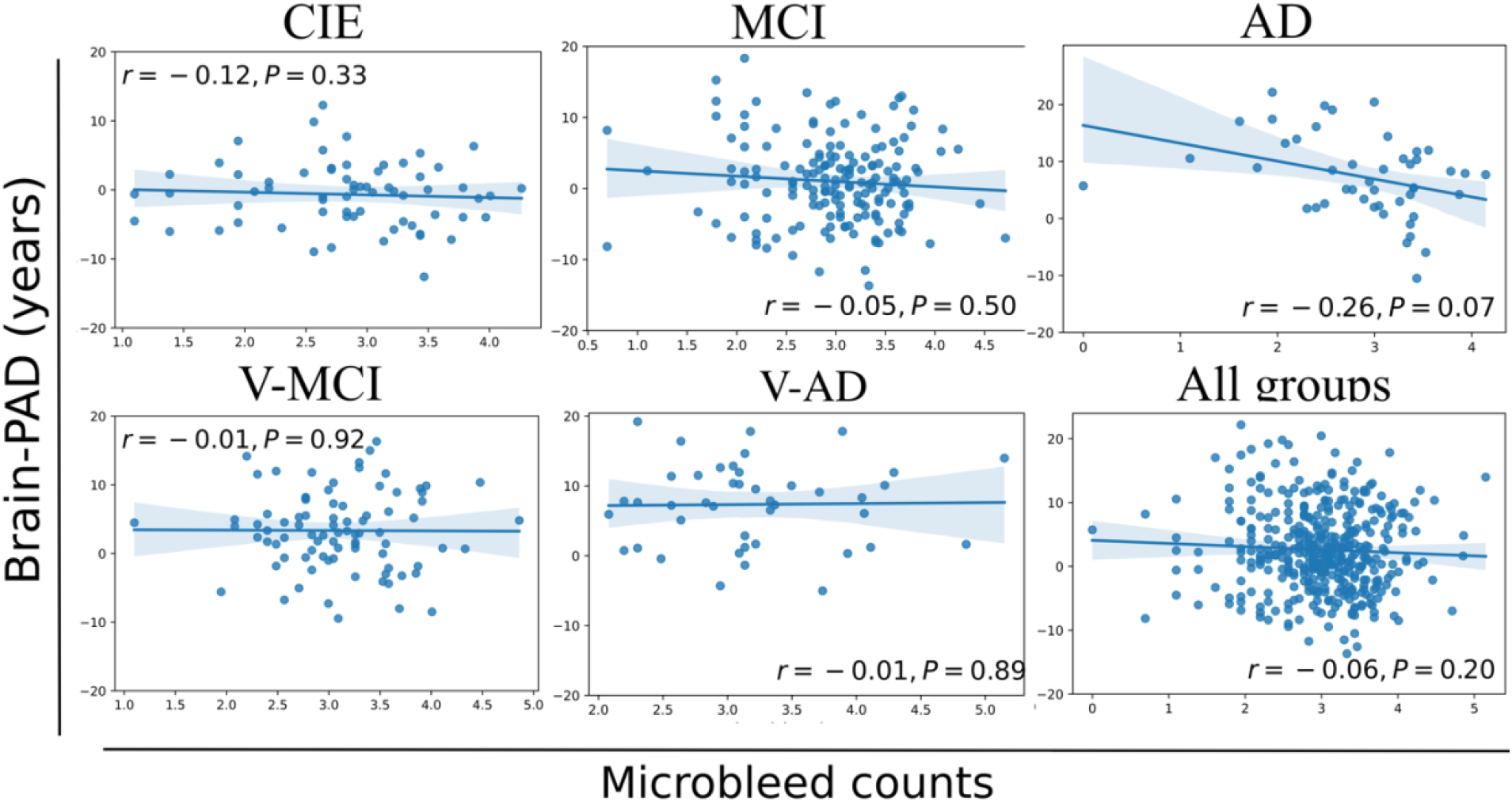
The association between brain-PAD values and microbleed counts in each cohort, as well as in all cohorts. The correlation analysis was conducted using a partial correlation test while controlling for age and sex. Microbleed counts were log-transformed.

## 4. DISCUSSION

In this report, we explored the association between brain-PAD and cerebrovascular lesion load, including WMHs and microbleeds, in aging and across the spectrum of cognitive impairment due to AD and cerebrovascular etiologies. All groups showed significantly increased brain-PAD values, with the V-AD group demonstrating the highest mean brain-PAD of +7.61 years. Our study found a mean brain-PAD of +7.23 years in individuals with AD, which aligns with previous research reporting a similar increase of +5 years in brain-PAD among AD patients [10]. However, we observed a lower mean brain-PAD in our MCI cohort (i.e., 0.98 ± 5.75years; Table 1) compared to the existing literature [10].

This discrepancy could potentially be explained by the fact that unlike prior studies [10], we distinguished V-MCI patients from those with MCI, which may suggest more serious MCI cases. To the best of our knowledge, this is the first study which explored brain age among V-MCI and V-AD participants as well, in the same cohort and using a similar imaging protocol. We observed a significantly higher brain-PAD in V-MCI than MCI (mean difference = 2.5 years, *P* < 0.001, ANCOVA; Fig 1B). However, this difference was not significant between AD and V-AD although V-AD showed a higher brain-PAD (mean difference = 0.5 years, *P* > 0.05, ANCOVA; Fig 1B). Further study in the area of brain age estimation and neuroimaging are required to confirm this finding.

All participant categories showed a significant WMH load on T2w-FLAIR images (Fig 2A). As expected, WMH load in vascular cohorts (i.e., V-MCI, V-AD) were significantly higher than non-vascular cohorts (i.e., CIE, MCI and AD; Fig 2B). This finding is in line with other studies which investigated WMH in MCI and AD [32, 33]. However, we did not observe a significant difference between V-MCI and V-AD (*P* > 0.05, ANCOVA; Fig 1B).

Our results revealed a statistically significant and positive correlation (r = 0.36, *P* < 0.001) between brain-PAD and WMH loads when all groups were combined, suggesting that there is an actual relationship between these two variables rather than one caused by chance. When we tested the correlation between brain-PAD and WMH loads within each group, only the MCI cohort showed a significant association between the two variables, whereas the other cohorts did not. The possible explanation is that the sample size within each cohort, except for the MCI cohort, was relatively small compared to the total sample size of all subjects. The association between brain-PAD and WMH loads in the CIE, V-MCI and V-AD cohorts were close to the level of significance with a p-value of 0.06-0.08 and it is likely that a larger sample size within each group would have resulted in significant correlations. Therefore, this result should be taken into account in clinical settings. To gain a better understanding of the relationship between Brain-PAD and WMH loads in AD subgroups, larger sample sizes are required in future studies.

Based on the literature [16], our first hypothesis was that a heavy WMH load can increase brain age in cognitively healthy older adults. We observed a positive relationship between brain-PAD and WMH in the CIE cohort, however, it did not reach a significant level (r = 0.23, *P* = 0.06; Fig 4), thus negating our initial hypothesis. However, this result does suggest a noteworthy trend or potential relationship between brain-PAD and WMH in the CIE cohort. A positive association between brain-PAD and WMH in the CIE cohort is in agreement with other studies documenting how a high WMH burden coincides with accelerated brain ageing and grey matter atrophy [16]. These findings suggest that WMH should be considered as a potential risk factor for accelerated brain aging among cognitively healthy older adults, as it may increase the likelihood of converting to MCI within this population [19]. We also hypothesized that WMH loads are associated with increasing brain-PAD in the context of AD. With a significant correlation in the MCI cohort and very close to being significant in other AD subgroups, except for the V-AD cohort (Fig 4), this hypothesis was confirmed. It is noteworthy that, even though a P-value of 0.06-0.08 may not be statistically significant, it could still be of clinical or practical importance. In spite of the fact that these links may differ based on the diagnosis, these findings indicate the importance of treatment and prevention strategies for vascular risk factors (e.g., lifestyle changes, anti-hypertensive medications, lipid-lowering treatments, blood sugar management, and exercise), which might be able to slow down the progression of cerebrovascular lesions and delay the effect on cortical thickness. Future research studies may aim to assess the efficiency of different WMH treatments and prevention strategies in the area of brain ageing.

Consistent with expectations, all categorical cohorts exhibited a notable presence of microbleeds (Fig. 3A), but no statistically significant differences were observed among the test cohorts (Fig. 3 B). Negative but insignificant associations between Brain-PAD and microbleed counts were detected in all cohorts (Fig. 5). However, it is worth noting that these correlations were derived as a result of applying a log-transformed operation on microbleed loads. Taken together, it can be inferred that the presence of WMH loads has the potential to significantly accelerate brain aging not only in the context of AD but also among cognitively healthy older adults. However, the impact of microbleed loads may not be as significant.

Evidence has shown that younger AD patients tend to exhibit higher levels of brain-PAD compared to older individuals with AD a higher brain-PAD than those who are older [6, 10]. In our study, we observed differences in the average age among our test groups (Table 1). While we accounted for the impact of age in our statistical analysis, it remains vital to acknowledge and address this point to prevent biases in interpreting the results, especially when comparing different clinical groups.

Finally, we highlight the fact that our brain age prediction model was created using data from multiple sites and scanners, including ADNI, OASIS, BAI, and ARWIBO, thus highlighting the generalizability of our results. In MRI pre-processing stage, we employed a validated software, specifically *FreeSurfer*, renowned for its suitability in multi-center and multi-scanner studies [34]. The test datasets were entirely independent from the training dataset. To ensure the separation of brain features extracted by *FreeSurfer* from WMH [35], only cortical brain features were used in our brain age prediction model.

## 5. Conclusion

This study aimed to investigate the relationship between brain-PAD and the cerebrovascular lesion loads in the context of aging and various forms of AD. In pursuit of this objective, we obtained the brain-PAD values, WMH loads and microbleed counts using a larger set of MRI scans obtained from 70 CIE, 173 individuals with MCI, 88 V-MCI, 50 with AD, and 47 with V-AD. Our results revealed a substantial link between brain-PAD and WMH loads, indicating a strong association between WMHs and accelerated brain aging, resulting in an older-appearing brain. Conversely, there was no significant correlation observed between brain-PAD and microbleed loads. Taken together, these findings provide comprehensive insights into the impact of WMHs on brain aging and underscore the importance of prevention and treatment strategies targeting WMHs.

## Data Availability

All data produced are available online at
https://adni.loni.usc.edu/data-samples/access-data/

## ACKNOWLEDGMENTS

Data used in this article were obtained from various datasets, including the Comprehensive Assessment of Neurodegeneration and Dementia (COMPASS-ND) cohort of the Canadian Consortium for Neurodegeneration and Aging (CCNA), Alzheimer’s Disease Neuroimaging Initiative (ADNI), Alzheimer’s Disease Repository Without Borders (ARWIBO), and The Open Access Series of Imaging Studies (OASIS). We express our sincere gratitude to all the principal investigators involved in the collection of these datasets, who collected these datasets and agreed to let them accessible. We also extend our appreciation to the esteemed participants who volunteered to be a part of these datasets, as their participation and cooperation were essential in advancing our understanding in this field.

- CCNA is supported by a grant from the Canadian Institutes of Health Research with funding from several partners.
- ADNI which was funded by National Institutes of Health (Grant U01 AG024904) and DOD ADNI (Department of Defense award number W81XWH-12-2-0012), the National Institute on Aging, the National Institute of Biomedical Imaging and Bioengineering, and through generous contributions from the following: AbbVie, Alzheimer’s Association; Alzheimer’s Drug Discovery Foundation; Araclon Biotech; BioClinica, Inc.; Biogen; Bristol-Myers Squibb Company; CereSpir, Inc.; Cogstate; Eisai Inc.; Elan Pharmaceuticals, Inc.; Eli Lilly and Company; EuroImmun; F. Hoffmann-La Roche Ltd and its affiliated company Genentech, Inc.; Fujirebio; GE Healthcare; IXICO Ltd.; Janssen Alzheimer Immunotherapy Research & Development, LLC.; Johnson & Johnson Pharmaceutical Research & Development LLC.; Lumosity; Lundbeck; Merck & Co., Inc.; Meso Scale Diagnostics, LLC.; NeuroRx Research; Neurotrack Technologies; Novartis Pharmaceuticals Corporation; Pfizer Inc.; Piramal Imaging; Servier; Takeda Pharmaceutical Company; and Transition Therapeutics. The Canadian Institutes of Health Research is providing funds to support ADNI clinical sites in Canada. Private sector contributions are facilitated by the Foundation for the National Institutes of Health (www.fnih.org). The grantee organization is the Northern California Institute for Research and Education, and the study is coordinated by the Alzheimer’s Therapeutic Research Institute at the University of Southern California. ADNI data are disseminated by the Laboratory for Neuro Imaging at the University of Southern California.
- ARWIBO was obtained from NeuGRID4You initiative (www.neugrid4you.eu) funded by the European Commission (FP7/2007-2013) under grant agreement no.283562.
- OASIS was supported by grants P50 AG05681, P01 AG03991, R01 AG021910, P50 MH071616, U24 RR021382, R01 MH56584).

## COMPETING INTERESTS

S.D. is co-founder and officer of True Positive Medical Devices, inc., All other authors declare no competing interests.

## Notes

### Funding Statement

This study did not receive any funding

### Author Declarations

we used data from public datasets. Ethical agreement Ethical agreements were obtained at all respective sites. Written informed consent was obtained from all participants.

## REFERENCES

[1] I. Beheshti, H. Demirel, H. Matsuda, and A. s. D. N. Initiative, “Classification of Alzheimer’s disease and prediction of mild cognitive impairment-to-Alzheimer’s conversion from structural magnetic resource imaging using feature ranking and a genetic algorithm,” (in eng), Comput Biol Med, vol. 83, pp. 109–119, 04 2017, doi: 10.1016/j.compbiomed.2017.02.011.

[2] K. Franke and C. Gaser, “Ten years of brainage as a neuroimaging biomarker of brain aging: what insights have we gained?,” (in eng), Front Neurol, vol. 10, p. 789, 2019, doi: 10.3389/fneur.2019.00789.

[3] D. Sone and I. Beheshti, “Neuroimaging-based brain age estimation: a promising personalized biomarker in neuropsychiatry,” Journal of Personalized Medicine, vol. 12, no. 11, p. 1850, 2022.

[4] S. Mishra, I. Beheshti, and P. Khanna, “A Review of Neuroimaging-driven Brain Age Estimation for identification of Brain Disorders and Health Conditions,” IEEE Reviews in Biomedical Engineering, 2021.

[5] H. R. Pardoe and R. Kuzniecky, “NAPR: a Cloud-Based Framework for Neuroanatomical Age Prediction,” Neuroinformatics, vol. 16, no. 1, pp. 43–49, Jan 2018, doi: 10.1007/s12021-017-9346-9.

[6] I. Beheshti, S. Nugent, O. Potvin, and S. Duchesne, “Disappearing metabolic youthfulness in the cognitively impaired female brain,” (in eng), Neurobiol Aging, vol. 101, pp. 224–229, 05 2021, doi: 10.1016/j.neurobiolaging.2021.01.026.

[7] C. Gaser, K. Franke, S. Klöppel, N. Koutsouleris, H. Sauer, and A. s. D. N. Initiative, “BrainAGE in Mild Cognitive Impaired Patients: Predicting the Conversion to Alzheimer’s Disease,” (in eng), PLoS One, vol. 8, no. 6, p. e67346, 2013, doi: 10.1371/journal.pone.0067346.

[8] K. Franke and C. Gaser, “Longitudinal changes in individual BrainAGE in healthy aging, mild cognitive impairment, and Alzheimer’s disease,” GeroPsych, 2012.

[9] D. Sone et al., “Neuroimaging-derived brain age is associated with life satisfaction in cognitively unimpaired elderly: A community-based study,” (in eng), Transl Psychiatry, vol. 12, no. 1, p. 25, 01 20 2022, doi: 10.1038/s41398-022-01793-5.

[10] I. Beheshti, N. Maikusa, and H. Matsuda, “The association between “Brain-Age Score”(BAS) and traditional neuropsychological screening tools in Alzheimer’s disease,” Brain and Behavior, vol. 8, no. 8, p. e01020, 2018.

[11] M. Habes et al., “The brain chart of aging: Machine-learning analytics reveals links between brain aging, white matter disease, amyloid burden, and cognition in the iSTAGING consortium of 10,216 harmonized MR scans,” Alzheimer’s & Dementia, vol. 17, no. 1, pp. 89–102, 2021.

[12] P. A. Boyle, L. Yu, R. S. Wilson, S. E. Leurgans, J. A. Schneider, and D. A. Bennett, “Person-specific contribution of neuropathologies to cognitive loss in old age,” Ann Neurol, Dec 15 2017, doi: 10.1002/ana.25123.

[13] P. A. Boyle et al., “Varied effects of age-related neuropathologies on the trajectory of late life cognitive decline,” (in eng), Brain, vol. 140, no. 3, pp. 804–812, Mar 01 2017, doi: 10.1093/brain/aww341.

[14] M. Dadar et al., “White Matter Hyperintensity Distribution Differences in Aging and Neurodegenerative Disease Cohorts,” bioRxiv, 2021.

[15] E. E. Smith et al., “Prevention of Stroke in Patients With Silent Cerebrovascular Disease: A Scientific Statement for Healthcare Professionals From the American Heart Association/American Stroke Association,” (in eng), Stroke, vol. 48, no. 2, pp. e44–e71, Feb 2017, doi: 10.1161/STR.0000000000000116.

[16] M. Habes et al., “White matter hyperintensities and imaging patterns of brain ageing in the general population,” (in eng), Brain, vol. 139, no. Pt 4, pp. 1164–79, Apr 2016, doi: 10.1093/brain/aww008.

[17] S. W. Seo et al., “Cardiovascular risk factors cause cortical thinning in cognitively impaired patients: relationships among cardiovascular risk factors, white matter hyperintensities, and cortical atrophy,” Alzheimer Disease & Associated Disorders, vol. 26, no. 2, pp. 106–112, 2012.

[18] S. W. Seo et al., “Cortical thinning related to periventricular and deep white matter hyperintensities,” Neurobiology of aging, vol. 33, no. 7, pp. 1156–1167. e1, 2012.

[19] A. Soldan et al., “White matter hyperintensities and CSF Alzheimer disease biomarkers in preclinical Alzheimer disease,” Neurology, vol. 94, no. 9, pp. e950–e960, 2020.

[20] S. M. Greenberg et al., “Cerebral microbleeds: a guide to detection and interpretation,” (in eng), Lancet Neurol, vol. 8, no. 2, pp. 165–74, Feb 2009, doi: 10.1016/S1474-4422(09)70013-4.

[21] L. Puy et al., “Cerebral microbleeds: from depiction to interpretation,” (in eng), J Neurol Neurosurg Psychiatry, Feb 09 2021, doi: 10.1136/jnnp-2020-323951.

[22] H. Chertkow et al., “The Comprehensive Assessment of Neurodegeneration and Dementia: Canadian Cohort Study,” Can J Neurol Sci, vol. 46, no. 5, pp. 499–511, Sep 2019, doi: 10.1017/cjn.2019.27.

[23] P. B. Gorelick et al., “Vascular contributions to cognitive impairment and dementia: a statement for healthcare professionals from the american heart association/american stroke association,” Stroke, vol. 42, no. 9, pp. 2672–713, Sep 2011, doi: 10.1161/STR.0b013e3182299496.

[24] P. Sachdev et al., “Diagnostic criteria for vascular cognitive disorders: a VASCOG statement,” Alzheimer Dis Assoc Disord, vol. 28, no. 3, pp. 206–18, Jul-Sep 2014, doi: 10.1097/WAD.0000000000000034.

[25] M. S. Albert et al., “The diagnosis of mild cognitive impairment due to Alzheimer’s disease: recommendations from the National Institute on Aging-Alzheimer’s Association workgroups on diagnostic guidelines for Alzheimer’s disease,” (in eng), Alzheimer’s & dementia : the journal of the Alzheimer’s Association, Research Support, Non-U.S. Gov’t vol. 7, no. 3, pp. 270–9, May 2011, doi: 10.1016/j.jalz.2011.03.008.

[26] G. M. McKhann et al., “The diagnosis of dementia due to Alzheimer’s disease: recommendations from the National Institute on Aging-Alzheimer’s Association workgroups on diagnostic guidelines for Alzheimer’s disease,” (in eng), Alzheimer’s & dementia, Consensus Development Conference, NIH Research Support, Non-U.S. Gov’t vol. 7, no. 3, pp. 263–9, May 2011, doi: 10.1016/j.jalz.2011.03.005.

[27] S. Duchesne et al., “The Canadian Dementia Imaging Protocol: Harmonizing National Cohorts,” (in eng), J Magn Reson Imaging, vol. 49, no. 2, pp. 456–465, Feb 2019, doi: 10.1002/jmri.26197.

[28] A. Klein and J. Tourville, “101 labeled brain images and a consistent human cortical labeling protocol,” (in eng), Front Neurosci, vol. 6, p. 171, 2012, doi: 10.3389/fnins.2012.00171.

[29] M. Dadar et al., “Performance comparison of 10 different classification techniques in segmenting white matter hyperintensities in aging,” (in eng), Neuroimage, vol. 157, pp. 233–249, 08 15 2017, doi: 10.1016/j.neuroimage.2017.06.009.

[30] M. Dadar, M. Zhernovaia, S. Mahmoud, R. Camicioli, J. Maranzano, and S. Duchesne, “Using Transfer Learning for Automated Microbleed Segmentation,” bioRxiv, p. 2022.05. 02.490283, 2022.

[31] I. Beheshti, S. Nugent, O. Potvin, and S. Duchesne, “Bias-adjustment in neuroimaging-based brain age frameworks: A robust scheme,” (in eng), Neuroimage Clin, vol. 24, p. 102063, 2019, doi: 10.1016/j.nicl.2019.102063.

[32] P. Desmarais et al., “White matter hyperintensities in autopsy-confirmed frontotemporal lobar degeneration and Alzheimer’s disease,” (in eng), Alzheimers Res Ther, vol. 13, no. 1, p. 129, 07 13 2021, doi: 10.1186/s13195-021-00869-6.

[33] G. Tosto, M. E. Zimmerman, O. T. Carmichael, A. M. Brickman, and A. s. D. N. Initiative, “Predicting aggressive decline in mild cognitive impairment: the importance of white matter hyperintensities,” (in eng), JAMA Neurol, vol. 71, no. 7, pp. 872–7, Jul 01 2014, doi: 10.1001/jamaneurol.2014.667.

[34] M. Dadar, S. Duchesne, and C. G. a. t. C.-Q. Group, “Reliability assessment of tissue classification algorithms for multi-center and multi-scanner data,” (in eng), Neuroimage, vol. 217, p. 116928, Aug 15 2020, doi: 10.1016/j.neuroimage.2020.116928.

[35] M. Dadar, O. Potvin, R. Camicioli, S. Duchesne, and A. s. D. N. Initiative, “Beware of white matter hyperintensities causing systematic errors in FreeSurfer gray matter segmentations!,” (in eng), Hum Brain Mapp, vol. 42, no. 9, pp. 2734–2745, Jun 15 2021, doi: 10.1002/hbm.25398.

